# Adapting the daily inventory of stressful events for SMS delivery: Recommendations for research

**DOI:** 10.1101/2025.04.15.25325757

**Authors:** Emily Hales, Nathan P. Helsabeck, Ana Carolina Wong, Kathy D. Wright

## Abstract

The everyday impacts of stressful experiences lead to an increased risk of heart disease, stroke, and mental illness. The Daily Inventory of Stressful Events (DISE) is a widely used measure of daily stressors. The DISE’s popularity is based on its accepted validity and the detailed information it provides researchers. However, the DISE relies on interviews conducted one-on-one with participants over several days and, as such, presents a high burden to both participants and researchers. Thus, many researchers have opted to simplify the measure by substituting the interview with a battery of survey questions. For a feasibility study addressing dementia caregiver stress and self-care, this paper aims to examine the lessons learned from an adaptation delivered via phone text messages. While the adaptation was used to reduce participant and researcher burden, it still provides high-quality information about the variability in daily stressors experienced by participants. However, the quality of the data collected was hindered by several design choices made in the adaptions that limited the utility of the data. As such, this paper provides recommendations for enhancing the utility of mobile phone text-based delivery of both the DISE and other measures and highlights the need for further validity work on such adaptations.

## Introduction

The daily insults of stressful experiences lead to an increased risk of heart disease, stroke, and mental illness (1,2). In general, the lost revenue for American companies has been attributed to stress-related factors such as absenteeism, lower productivity, and worker turnover, which is over 200 million U.S. dollars annually (3). For dementia caregivers, 70% report that activities such as coordinating care are their top stressors (3). Stress is a contributor to a myriad of both psychological and physical health problems for dementia caregivers, such as hypertension, anxiety, depression, and financial loss of wages (4). As such, many measures have been developed to help understand both the causes of stress and individuals’ reactions to that stress. Among the most widely used and trusted measures of daily stressors is the Daily Inventory of Stressful Events (DISE) (5,6).

The DISE was developed as a measure of the prevalence of daily stressful interactions, the severity of those interactions, and the perceived threats posed by those interactions (6,7). A strength of the measure is that it employs one-to-one interviews by phone or in person rather than relying on a self-administered checklist or inventory (5,6). To administer the DISE, an interviewer asks seven stem questions (e.g., “Did you have an argument or disagreement with anyone since this time yesterday?”). Respondents who reply “yes” are asked a short series of follow-up questions about the severity and perception of threats. This interview process allows the interviewer to follow up and probe as needed.

Yet, conducting interviews is both time-consuming and costly in applied research settings. As such, over the past two decades, researchers have adapted both the quantification and the delivery of the DISE. Regarding quantification, a common practice has been to examine only the initial stem questions to quantify the number of stressful events that occurred on a given day (8–12). In some cases, specific items were selected for use instead of the full set of seven (8,13); on at least one occasion, stressful events were categorized into interpersonal and non-interpersonal events (11). When only stem questions are used, sum scores are generally calculated based on a score of one for an event that occurs and is examined for variability over the study. Another adaption type involves examining the frequency and severity of stressful events. In these cases, the stem question is asked, and if respondents answer affirmatively, a follow-up question regarding the event’s severity is asked (14,15).

To make the DISE less time-consuming, the DISE has been adapted to facilitate easier data collection, reduce participant burden, and increase the amount of data that can be collected. To accomplish this, researchers have used smartphone surveys delivered by Short Message Service (SMS) text messaging (15,16), paper surveys (10), journals (5), or provided links for participants to complete surveys online (8,14). However, the commonality of these adaptations is that they remove interviewers from the process, thus removing one of the strengths of the DISE (5). To date, we have found no evidence of validity work being conducted to support these adaptations in the studies we reviewed.

The current study evaluates the delivery of the DISE via SMS during a pilot study for an intervention focused on reducing stress and improving self-care behaviors in family caregivers of people living with dementia (17,18). Because the focus is on the effectiveness of the delivery, we do not consider the outcomes of the participants or examine the differences between the treatment and control conditions. Rather, the purpose is to explore the extent to which SMS delivery of the DISE is feasible and appropriate for participants. Furthermore, where barriers to completion arise, we aim to utilize this information to develop a more accessible version for future work.

## Method

### Participants and Procedure

The study titled, “Addressing the double jeopardy of stress and hypertension among African American female caregivers of persons living with Alzheimer’s disease and related dementias,” was a randomized control trial funded by the National Institute on Aging. This study aimed to determine the feasibility, acceptability, and impact of a Mindfulness in Motion plus the Dietary Approaches to Stop Hypertension intervention for African American female caregivers with hypertension (17). The research also investigated the potential mediating effects of stress reactivity and stress resilience on self-care behaviors among the intervention, Alzheimer’s Association Caregiver education, and attention control groups. Approval for this study was granted buy The Ohio State University Behavioral and Social Sciences institutional review board (IRB; approval 2022B0031). All participants were provided written informed consent before their inclusion. Participants were recruited for the study in the Midwest through partnerships with community sites that had connections to caregivers of individuals living with dementia, such as the African American Alzheimer’s and Wellness Association, as well as through advertising on ResearchMatch and social media sites (19). Consent was obtained for all participants and data was analyzed anonymously. Participants were recruited between February 1, 2023 and November 30, 2023.

Using SMS, the participants were asked to complete the DISE survey daily for seven days in a row at three different time points: baseline, 3 months post-intervention, and 9 months post-intervention. We refer to variance within a seven-day administration as *within week* and variance between the three administrations as *between time points*.

We adapted the DISE as an SMS delivered survey using Mosio (20). Mosio is an automatic SMS software designed for researchers to send scheduled and automatic texts to participants whose responses are recorded. Branching expressions were built into each question based on participant responses. To accurately direct participants through the survey, predefined allowable responses were specified for each branching expression. The Mosio system evaluates each participant’s response to determine the next step in the survey. If a participant’s response did not meet the required conditions, Mosio sent an automatic error message and prompted participants to try again. We used the Mosio Storyline feature to schedule the DISE survey to be sent at 6 PM each day for one week. As an alternative, participants were also offered the opportunity to complete the DISE by scheduling a daily telephone call with a research assistant.

### Measures

The current study examines an adaptation of DISE—our adaptation is available in Supplemental Table 1. To explore the adaptation of the adapted DISE, we examined participant responses. Our definitions of these responses follow.

#### Valid Responses

Valid responses are defined as participant responses recognized by SMS to be within the accepted predetermined parameters. All other responses are considered invalid. Participants were informed that their invalid responses were rejected through an automatic error message.

#### Survey completion

A survey is considered complete if all relevant branching questions have an associated valid response. As this is a daily survey taken over the course of a week, a total of 7 completed surveys within week are possible per individual at each time point.

#### Survey frustration

Survey frustration was identified as a discontinued survey following an invalid user response.

#### Survey fatigue

Survey fatigue, or an increasing disengagement as the survey progressed, was measured within week by the higher percentage of incomplete surveys later in the week compared to earlier in the week.

#### Question type

Questions were categorized into the following types: choose all that apply (1 question), single-choice exclusive (10 questions), single-choice nonexclusive/multiple potential options (4 questions), and time (1 question). Three questions were free-response and are not included in our analysis as any response was considered valid.

### Analysis Plan

To assess the feasibility and usability of SMS delivery for DISE, we utilized data accessed through the text message delivery system. We examined completion rates (both within week and between time points), the number of valid responses, and the rate of survey dropouts. Completion rates are reported as the number and percentage of surveys initiated and completed across all time points. Valid responses are evaluated in several ways: the percentage of invalid responses by question type, the number of attempts before a valid response is provided, and the number of invalid responses preceding the point of incompletion. Survey dropouts are reported as an outcome following invalid responses by question type. We examined dropout patterns, both generally and over time, in relation to question type, survey frustration, and survey fatigue. All analysis was conducted in SAS version 9.4.

## Results

Of the 28 individuals participating in our larger study, 24 participated in the DISE through SMS. Overall, 86.9% of SMS-delivered DISE surveys were completed, 4.5% were partially completed (responded to but not completed), and 8.6% received no response (see **Table 1**). Although we did not find a pattern of declining participation within week, participation dropped between time points.

**Table 1.** DISE SMS Response Rates.

|  | SMS Survey<br>Participants | Total surveys<br>sent out | Completed<br>surveys | Partially<br>completed<br>surveys | Surveys not<br>responded to |
| --- | --- | --- | --- | --- | --- |
| Timepoint | n | n | n (%) | n (%) | n (%) |
| 1 | 24 | 169* | 153 (90.5%) | 6 (3.6%) | 10 (5.9%) |
| 2 | 20 | 140 | 121 (86.4%) | 7 (5.0%) | 12 (8.6%) |
| 3 | 16 | 112 | 92 (82.1%) | 6 (5.4%) | 14 (12.5%) |
| Overall | 24 | 421 | 366 (86.9%) | 19 (4.5%) | 36 (8.6%) |
| *Note: One person was sent a survey eight times at time point 1; all others received seven surveys per time point.* |  |  |  |  |  |

Of the participant’s SMS responses, 97.4% were recognized as valid on the first attempt; however, this varied widely by question type. The question asking for the time of the stressful event (in the form of the hour, minute, and am/pm) had the highest rate of invalid responses on the first attempt (17.6%), with 7.1% of those dropping out, resulting in a 1.3% overall rate.

Single-choice questions with multiple, non-exclusive options (4 questions) had the highest overall dropout rate among respondents (2.0%), with 35.7% dropping out of the survey after encountering an invalid response on their first attempt. Questions with a single, exclusive choice (10 questions) were answered validly at first attempt 99.3% of the time with a 0.1% overall dropout rate; however, it should be noted that these questions were administered later in the survey, so they were only sent to individuals who correctly answered previous questions.

Overall, 14.5% of surveys were not completed after a SMS rejected response (see **Table 2**). Of questions receiving an initial invalid response, the median number of invalid responses preceding either a change to a valid response or a survey dropout was 1 (maximum = 3 for a change to a valid response; maximum = 7 for a survey dropout).

**Table 2.** SMS accepted Responses by Question Type.

| Question Type | First attempt |  | Outcome after invalid first attempt |  | Dropout, overall % |
| --- | --- | --- | --- | --- | --- |
|  | valid n (%) | invalid n (%) | valid n (%) | dropout n (%) |  |
| Choose all that apply | 376 (97.7%) | 9 (2.3%) | 8 (88.9%) | 1 (11.1%) | 0.3% |
| Single choice (multiple options) | 241 (94.5%) | 14 (5.5%) | 9 (64.3%) | 5 (35.7%) | 2.0% |
| Time | 131 (82.4%) | 28 (17.6%) | 26 (92.9%) | 2 (7.1%) | 1.3% |
| Single choice (exclusive) | 1535 (99.3%) | 11 (.7%) | 10 (90.9%) | 1 (9.1%) | 0.1% |
| Overall | 2283 (97.4%) | 62 (2.6%) | 53 (85.5%) | 9 (14.5%) | 0.4% |

We should note that some responses accepted by SMS as “valid” were logically invalid (e.g., conflicting options, Unicode characters accepted as numeric values, etc.). In contrast, others rejected as “invalid” were logically valid (e.g., responding with the text value instead of the corresponding numeric value). We also received automatic responses from users in “Focus” mode while driving. Although rare, these unanticipated issues, unique to text message delivery, contributed to user frustration and data loss.

## Discussion

Our SMS delivery of DISE resulted in high response rates for our population. This enabled us to incorporate DISE into our study, thereby improving our understanding of caregiver stress. Despite the challenges of adapting to SMS delivery from interview-based surveys, we had high rates of valid responses and high survey completion rates. Although SMS delivery fundamentally changes the nature of the survey, the adaptation is more accessible and enables us to analyze daily stress in a manner consistent with current literature. However, after reviewing the responses to our pilot study, we further revised our SMS DISE adaptation based on technical issues, analytic needs, and further testing. Below, we summarize how we addressed the unique challenges of adapting SMS delivery. For those who would like to capture a majority of the full DISE measure without incorporating daily phone interviews, we recommend making these improvements alongside SMS delivery to minimize errors, participant frustration, and missing data. The specific changes identified here may be useful for adapting survey instruments other than DISE for SMS delivery.

First, to improve data clarity and to both minimize participant frustration and missing responses, we recommend changing questions to single-choice exclusive whenever possible. Select-all-that-apply responses may confuse users and produce messy data that require significant cleaning. For example, in our revision, the occurrence of each type of stressor is asked in individual yes/no questions, rather than the original ‘select-all-that-apply’ format. Although many studies analyze only selective types of stressful interactions (8,13). We chose to keep all questions regarding interactions due to our study’s focus on stress.

Next, we recommend minimizing the number of options for single-choice, non-exclusive questions (i.e., multiple potential options that are not select-all-that-apply). For example, we collapsed the possible selections for “who” an argument was with from the 21 options originally identified in the interview guide to 6 categories. Making clarifications relevant to the study population can also be helpful. As we collapsed categories, we incorporated the option’ person with memory issues whom you help” and clarified “parent or grandparent, other than the care recipient,” since we are studying the stress of caregivers of individuals with dementia. Limiting text within a message and only offering choices relevant to the study may lessen the burden on respondents.

Third, we recommend modifying the validation criteria to reject responses containing non-numeric characters for consistency. This prevents accepting responses that include valid numbers but are not logically valid, such as an automatic reply including Unicode characters. Additionally, the automatic error message sent by SMS after an invalid participant response should clearly identify unacceptable responses. In addition to “Sorry, we do not recognize the value of that answer,” we have updated our error message to say, “Please try again and remember to respond with a single number that corresponds to one of the provided choices. ‘ Do not include special characters or words.” The automatic error message still included a phone number to call with questions. Accepting only numeric values for any SMS delivered survey will provide cleaner data and likely avoid confusion for participants.

Fourth, we recommend eliminating questions that cause validation issues and will not be used in the analysis. After reviewing common strategies for analyzing DISE, we removed the question regarding the time of day of the stressful event, which had the highest rate of invalid first attempts in our pilot study. In our case, the time of day was not of interest in the analysis, so removing it to simplify the survey made sense. In other surveys, it may be useful to consider eliminating items that are not relevant to the study or finding ways to simplify response choices for items that are of interest.

Finally, we recommend testing the SMS survey on phones with different cell phone providers before administering the survey. In our experience, one cell phone provider started blocking a message asking participants to identify with the “sex” of the person they cared for. In response, we changed “sex” to “gender,” which eliminated the issue.

Incorporating these changes improved our DISE adaptation. Pairing question simplification with a tightening of SMS automated delivery rules will help minimize participant frustration and maximize data cleanliness. For our study, this involved collapsing multiple choice options to those relevant to our population of interest, removing unnecessary questions, and clarifying acceptable responses. For others wishing to adapt their survey to SMS delivery, we recommend thoroughly testing the following: the branching logic against automated replies (e.g., Focus or Driving mode), special characters, and emojis; the delivery of texts to a variety of cell phone providers; the SMS delivery system’s automatic replies to different combinations of numeric and text responses. See Supplemental Table 2 for the questions, branching logic, and validation criteria for the updated SMS delivery adaptation of DISE.

## Conclusion

We recognize that adapting the delivery of a survey from interviews to SMS delivery affects the quality of the data collected, particularly when responses are automatically accepted or rejected. However, research participants increasingly opt for digital surveys over interviews due to the ease of administration (21–23). This preference holds for middle-aged and older adults(24). In our study, participants were also given the opportunity for daily phone calls, but only one preferred this method over SMS. Text message delivery minimizes the burden for both participants and research staff. Further, the logistical difficulty in collecting interview data combined with the high comparability data quality of digitally collected surveys make SMS a potentially powerful research tool (25). Overall, we found text message delivery of daily surveys to be useful, and we plan to continue using it for the daily inventory of stressful events.

## Data Availability

The data are available by request from the corresponding author.

## Acknowledgments

KDW is supported by the Robert Wood Johnson Foundation Harold Amos Medical Faculty Development Program.

