## Supplemental Table 1 for "Adapting the daily inventory of stressful events for SMS delivery: Recommendations for research"

**Supplemental Table 1. Pilot SMS-delivery Adaptation of the Daily Inventory of Stressful Events**

| **Q #** | **Question** | **Question Type** | **Mosio Branching Expression(s) and Accepted Responses** |
| --- | --- | --- | --- |
| **1** | Which of the following types of stressors have you experienced today (text back all the numbers that apply, for example "1, 2, 3"):  1) Argument or disagreement with anyone  2) Work or school related event  3) Home related event  4) Discrimination on the basis of race/sex/age  5) Close friend or relative event that was stressful for you  6) Anything else that people would consider stressful  7) None | Choose all that apply | Branching Expressions:  [This answer][equal to] [7], go to [Complete].  [This answer][regular expression] [1], go to [Q2].  [This answer][regular expression] [4], go to [Q4].  [This answer][regular expression] [5], go to [Q6].  [This answer][regular expression] [(2\|3\|6)], go to [Q8]. |
| **2** | If argument or disagreement...Who was it with (select the most stressful option):  1) Spouse or partner  2) Child or grandchild  3) Parent  4) Sibling  5) Other relative  6) Friend  7) Neighbor  8) Coworker or fellow student  9) Boss or teacher  10) Employee or supervisee  11) Other (specify)  12) Stranger  13) Religious group member  14) Self-help group  15) Client/customer/patient  16) Groups  17) Landlord or realtor  18) Family  19) Pets  20) Doctors/nurses/health professionals  21) Home related people | Single choice (multiple options) | Branching Expressions:  [This answer][regular expression][11], go o [Q3].  [Previous Answer][1][regular expression][4] AND [This answer][regular expression][^(\d\|1\d\|20\|1)$], go to [Q4].  [Previous Answer][1][regular expression][5] AND [This answer][regular expression][^(\d\|1\d\|20\|1)$], go to [Q6].  [This answer][regular expression][^(\d\|1\d\|20\|21)$], go to [Q8]. |
| **3** | Please specify other (argument or disagreement): |  | Branching Expressions:  [Previous Answer][1][regular expression][4], go to [Q4].  [Previous Answer][1][regular expression][5], go to [Q6].  [Default], go to [Q8]. |
| **4** | If discrimination...What was the basis for the discrimination you experienced (select the most stressful option):  1) Race  2) Sex  3) Age  4) Other (specify)  5) Something else (specify) | Single choice (multiple options) | Branching Expressions:  [This Answer][regular expression][^[45]$], go to [Q5].  [Previous Answer][1][regular expression][5] AND [This Answer][regular expression][^[123]$], go to [Q6].  [This Answer][regular expression][^[123]$], go to [Q8]. |
| **5** | Please specify other or something else (discrimination): |  | Branching Expressions:  [Previous Answer][1][regular expression][5], go to [Q6].  [Default], go to [Q8]. |
| **6** | If close friend or relative event...Who was it with (select the most stressful option):  1) Spouse or partner  2) Child or grandchild  3) Parent  4) Sibling  5) Other relative  6) Friend  7) Neighbor  8) Coworker or fellow student  9) Boss or teacher  10) Employee or supervisee  11) Home related people  12) Stranger  13) Religious group member  14) Self-help group  15) Client/customer/patient  16) Groups  17) Landlord or realtor  18) Family  19) Pets  20) Doctors/nurses/health professionals  21) Other (specify) | Single choice (multiple options) | Branching Expressions:  [This Answer][equal to][21], go to [Q7].  [This Answer][regular expression][^(\d\|1\d\|20)$], go to [Q8]. |
| **7** | Please specify other (close friend or relative event): |  | Branching Expressions:  [Default], go to [Q8]. |
| **8** | When did it [stressor] happen?  1) Yesterday  2) Today  3) I don't know | Single choice (multiple options) | Branching Expressions:  [This Answer][equal to][1], go to [Q9].  [This Answer][equal to][2], go to [Q9].  [This Answer][equal to][3], go to [Q10]. |
| **9** | What time of day did this happen (please record time in hours and minutes): For example, 5:30 PM | Time | Regular Expression:  /([1-9]:[0-5]\d\|1[012]:[0-5]\d)(\s\|)(am\|pm)/ |
| **10** | How stressful was this for you?  0) None at All  1) A little  2) Somewhat  3) Very | Single choice (exclusive) | User defined set of characters (case insensitive):  0123 |
| **11** | How much control did you have over the situation [stressor]?  0) None at All  1) A little  2) Somewhat  3) Very | Single choice (exclusive) | User-defined set of characters (case insensitive):  0123 |
| **12** | Is the issue [stressor] resolved?  1) Yes  0) No | Single choice (exclusive) | User-defined set of characters (case insensitive):  0123 |
| **13** | How much did it [stressor] disrupt your daily routine?  0) None at All  1) A little  2) Somewhat  3) Very | Single choice (exclusive) | User-defined set of characters (case insensitive):  0123 |
| **14** | How much did it [stressor] risk your financial situation?  0) None at All  1) A little  2) Somewhat  3) Very | Single choice (exclusive) | User-defined set of characters (case insensitive):  0123 |
| **15** | How much did it [stressor] risk the way you feel about yourself?  0) None at All  1) A little  2) Somewhat  3) Very | Single choice (exclusive) | User-defined set of characters (case insensitive):  0123 |
| **16** | How much did it [stressor] risk the way other people feel about you?  0) None at All  1) A little  2) Somewhat  3) Very | Single choice (exclusive) | User-defined set of characters (case insensitive):  0123 |
| **17** | How much did it [stressor] risk your physical health or safety?  0) None at All  1) A little  2) Somewhat  3) Very | Single choice (exclusive) | User-defined set of characters (case insensitive):  0123 |
| **18** | How much did it [stressor] risk the health or well-being of someone you care about?  0) None at All  1) A little  2) Somewhat  3) Very | Single choice (exclusive) | User-defined set of characters (case insensitive):  0123 |
| **19** | How much did it [stressor] risk your plans for the future?  0) None at All  1) A little  2) Somewhat  3) Very | Single choice (exclusive) | User-defined set of characters (case insensitive):  0123 |
