## Supplemental Table 2 for "Adapting the daily inventory of stressful events for SMS delivery: Recommendations for research"

**Supplemental Table 2. Updated SMS-delivery Adaptation of the Daily Inventory of Stressful Events**
Stem questions are highlighted in blue, and probes are in white.

| **Q #** | **Question** | **Mosio Branching Expression(s)** |
| --- | --- | --- |
| **1** | Did you have an argument or disagreement with anyone since this time yesterday?  1, Yes  2, No | [This answer] [equal to] [1], go to [Q8].  [This answer] [equal to] [2], go to [Q2]. |
| **8** | Think of the most stressful disagreement or argument you had since this time yesterday. Who was that with?  1) Person with memory issues whom you help  2) Spouse or partner  3) Child or grandchild  4) Parent or grandparent, other than care recipient  5) Doctors/nurses/health professionals  6) Other | [This answer] [regular expression] [(^1$\|^2$\|^3$\|^4$\|^5$\|^6$)], go to [Q9]. |
| **9** | How stressful was this for you?  0) None at All  1) A little  2) Somewhat  3) Very | [This answer] [regular expression] [(^0$\|^1$\|^2$\|^3$)], go to [Q2]. |
| **2** | Since this time yesterday, did anything happen that you could have argued about but you decided to let pass in order to avoid a disagreement?  1, Yes  2, No | [This answer] [equal to] [1], go to [Q10].  [This answer] [equal to] [2], go to [Q3]. |
| **10** | Think of the most stressful incident of this sort. Who was the person you decided not to argue with?  1) Person with memory issues whom you help  2) Spouse or partner  3) Child or grandchild  4) Parent or grandparent, other than care recipient  5) Doctors/nurses/health professionals  6) Other | [This answer] [regular expression] [(^1$\|^2$\|^3$\|^4$\|^5$\|^6$)], go to [Q11]. |
| **11** | How stressful was this for you?  0) None at All  1) A little  2) Somewhat  3) Very | [This answer] [regular expression] [(^0$\|^1$\|^2$\|^3$)], go to [Q3]. |
| **3** | Since this time yesterday, did anything happen at work (other than what you have already mentioned) that most people would consider stressful?  1, Yes  2, No | [This answer] [equal to] [1], go to [Q12].  [This answer] [equal to] [2], go to [Q4]. |
| **12** | How stressful was this for you?  0) None at All  1) A little  2) Somewhat  3) Very | [This answer] [regular expression] [(^0$\|^1$\|^2$\|^3$)], go to [Q4]. |
| **4** | Since this time yesterday, did anything happen at home (other than what you have already mentioned) that most people would consider stressful?  1, Yes  2, No | [This answer] [equal to] [1], go to [Q13].  [This answer] [equal to] [2], go to [Q5]. |
| **13** | How stressful was this for you?  0) None at All  1) A little  2) Somewhat  3) Very | [This answer] [regular expression] [(^0$\|^1$\|^2$\|^3$)], go to [Q5]. |
| **5** | Many people experience discrimination on the basis of such things as race, gender, body size, or age. Did anything like this happen to you since this time yesterday?  1, Yes  2, No | [This answer] [equal to] [1], go to [Q14].  [This answer] [equal to] [2], go to [Q6]. |
| **14** | Think of the most stressful incident of this sort. What was the basis for the discrimination you experienced—your race, gender, body size, age, or something else?  1) Race  2) Gender  3) Age  4) Body size  5) Other | [This answer] [regular expression] [(^1$\|^2$\|^3$\|^4$\|^5$)], go to [Q15]. |
| **15** | How stressful was this for you?  0) None at All  1) A little  2) Somewhat  3) Very | [This answer] [regular expression] [(^0$\|^1$\|^2$\|^3$)], go to [Q6]. |
| **6** | Since this time yesterday, did anything happen to a close friend or relative (other than what you have already mentioned) that turned out to be stressful for you?  1, Yes  2, No | [This answer] [equal to] [1], go to [Q16].  [This answer] [equal to] [2], go to [Q7]. |
| **16** | Think of the most stressful incident of this sort. Who did this happen to?  1) Person with memory issues whom you help  2) Spouse or partner  3) Child or grandchild  4) Parent or grandparent, other than care recipient  5) Doctors/nurses/health professionals  6) Other | [This answer] [regular expression] [(^1$\|^2$\|^3$\|^4$\|^5$\|^6$)], go to [Q17]. |
| **17** | How stressful was this for you?  0) None at All  1) A little  2) Somewhat  3) Very | [This answer] [regular expression] [(^0$\|^1$\|^2$\|^3$)], go to [Q7]. |
| **7** | Did anything else happen to you since this time yesterday that most people would consider stressful?  1, Yes  2, No | [This answer] [equal to] [1], go to [Q18].  [This answer] [equal to] [2], go to [Complete]. |
| **18** | How stressful was this for you?  0) None at All  1) A little  2) Somewhat  3) Very | [This answer] [regular expression] [(^0$\|^1$\|^2$\|^3$)], go to [Complete]. |
